# Circulating long noncoding RNA PDE4DIPP6: A novel biomarker for improving the clinical management of acute coronary syndrome

**DOI:** 10.1101/2024.01.30.24302038

**Authors:** Pia F. Koch, María C. García-Hidalgo, Josephine Labus, Moritz Biener, Thomas Thum, David de Gonzalo-Calvo, Christian Bär

## Abstract

**Aim:** Long noncoding RNAs (lncRNAs) have emerged as promising diagnostic biomarkers owing to their dynamic regulation in response to pathological conditions and their detection in clinically relevant samples. Here, we explored the utility of the cardiac expressed and plasma detectable lncRNA PDE4DIPP6 as a biomarker for acute coronary syndrome (ACS). The final goal was to improve the diagnostic efficacy of state-of-the-art tests, particularly the high-sensitivity cardiac troponin assay (hs-cTnT).

**Methods:** The study enrolled individuals presenting with suspected ACS at the emergency department (ED). LncRNA quantification was performed in plasma samples using RT-qPCR. Discriminatory performance was evaluated by calculating the Area Under the Curve (AUC). Reclassification metrics, including the Integrated Discrimination Improvement (IDI) and Net Reclassification Improvement (NRI) indexes, were employed to assess enhancements in diagnostic accuracy.

**Results:** The sample comprised 252 patients, 50.8% were diagnosed with ACS and 13.9% with Non-ST Segment Elevation Myocardial Infarction (NSTEMI). Elevated levels of PDE4DIPP6 were observed in ACS patients compared to non-ACS subjects. There was no significant correlation between lncRNA and hs-cTnT levels (rho=0.071), and no association between PDE4DIPP6 levels and potential confounding factors was observed. The inclusion of PDE4DIPP6 on top of troponin T significantly enhanced the discrimination and classification of ACS patients reflected in an improved AUC of 0.734, an IDI of 0.066 and NRI of 0.471. Similarly, elevated levels of the lncRNA were observed in NSTEMI patients compared to ACS patients without NSTEMI. Consistent with previous findings, the addition of PDE4DIPP6 to hs-cTnT improved the discrimination and classification of patients, evident in an increased AUC from 0.859 to 0.944, IDI of 0.237, and NRI of 0.658.

**Conclusion:** PDE4DIPP6 offers additional diagnostic insights beyond hs-cTnT, suggesting its potential to improve the clinical management of patients with ACS.

## Introduction

Acute coronary syndrome (ACS) encompasses a spectrum of symptoms, including chest pain, nausea, and dyspnea, indicative of acute myocardial ischemia.^1^ Clinical categorization of ACS includes unstable angina pectoris (uAP), ST-segment elevation myocardial infarction (STEMI) and Non-STEMI (NSTEMI).^2^ While uAP was traditionally defined as ACS without an elevation in cardiac enzymes,^3^ this view has been changed since the introduction of novel high-sensitivity cardiac troponin assay (hs-cTnT) assays.^4^ The hs-assays can detect cTnT elevations at concentrations 10–100-fold lower compared to conventional assays,^5^ resulting in improved acute myocardial infarction (AMI) detection and identification of smaller infarcts previously misdiagnosed as uAP. ^6^ However, the high sensitivity compromises clinical specificity, as cTnT release can occur in non-ACS pathologies.^7^ Consequently, supplementary parameters, in addition to hs-cTnT, are essential to enhance specificity for AMI diagnosis.^8^

Noncoding RNAs (ncRNAs) represent a rich but poorly investigated reservoir of biomarkers for various diseases, including heart conditions.^9^ ncRNAs, actively secreted through extracellular vesicles or released passively from dying cells, such as cardiomyocytes during ischemic episodes, can be readily identified in patient plasma or serum.^10–12^ While numerous studies have investigated small ncRNAs, particularly microRNAs, supporting their potential as biomarkers for managing cardiovascular conditions,^13,14^ there is limited data on long noncoding RNAs (lncRNAs) in this context. Given that lncRNAs exhibit dynamic regulation under stress or disease conditions, these transcripts emerge as a novel source for diagnostic, prognostic and predictive markers.^15^ A notable example is the circulating lncRNA LIPCAR, which is predictive for heart failure outcomes in patients with myocardial infarction^16^, particularly among elderly heart failure patients without chronic kidney disease.^17^

The current study aimed to investigate the potential of a novel plasma detectable and cardiac expressed lncRNA candidate as a biomarker in ACS and to test whether its quantification can improve the diagnostic efficacy of concurrent tests, with a particular focus on hs-cTnT.

## Methods

### Study population

The study design and characteristics of the whole study population including the diagnostic workup have been previously detailed (clinical trial identifier: NCT02116153). The study population included a subcohort of patients presenting between August 2014 and April 2017 with symptoms suggestive of an ACS to the emergency department (ED) of the Department of Cardiology, University Hospital Heidelberg, as described by our group.^18,19^ The study’s inclusion criteria encompassed individuals aged 18 years and above who provided written informed consent. Excluded from participation were individuals diagnosed with STEMI. Additionally, patients presenting with acute heart failure were not considered if the likelihood of acute myocardial ischemia was low or if there was the presence of tachycardias, pacemaker dysfunction or an Implantable Cardioverter-Defibrillator (ICD) alarm, coupled with a low pre-test probability of coronary artery disease. Furthermore, individuals presenting with conditions clearly unrelated to ACS, such as traumatic chest pain, pneumonia, chronic obstructive pulmonary disease (COPD), asthma, gastrointestinal disease, urinary tract infection, or those with unspecific symptoms, were also excluded from enrollment. Ultimately, this substudy includes 252 randomly selected patients available for lncRNA analysis. The median values of all clinical characteristics closely mirror those of the original cohort, as indicated in a previous study.^19^

The research received approval from the ethics committee of the University of Heidelberg and adhered to the principles outlined in the Declaration of Helsinki. Written informed consent was obtained from each participating patient.

### Cardiac troponin measurement

High-sensitivity cardiac troponin T (Hs-cTnT) levels were assessed according to the standard protocol. Plasma samples collected at admission were processed in the central laboratory of the University Hospital of Heidelberg. The measurement employed the hs-cTnT assay on a COBAS E411 (Roche Diagnostics, Mannheim, Germany), as previously described.^18,19^

### Circulating lncRNA selection, isolation and quantification

Phosphodiesterase 4D Interacting Protein Pseudogene 6 (PDE4DIPP6) is an intergenic lncRNA of 1485 nt in length located in chromosome 1. Our prior investigations have confirmed the expression of PDE4DIPP6 in both human induced pluripotent stem cell-derived cardiomyocytes (hiPSC-CM) and human cardiac fibroblasts (HCF) **(Supplemental Figure S1)**. PDE4DIPP6 has also been detected in different human samples, including heart, artery and whole blood, and cardiac cells [the Genotype-Tissue Expression (GTEx) Portal, (https://www.gtexportal.org/home/)]. The lncRNA has also been detected in circulating exosomes (GSE159657). Furthermore, PDE4DIPP6 is a pseudogene of PDE4DIP, a protein previously linked to cardiovascular diseases.^20^ Therefore, we speculated that PDE4DIPP6 would constitute a candidate biomarker in the context of ACS.

For the analysis of PDE4DIPP6, Ethylenediaminetetraacetic acid (EDTA) plasma samples were obtained during a routine blood draw at the time of presentation. Following the blood draw, plasma isolation was promptly conducted through centrifugation, with the separated plasma stored at −80°C. The centrifugation process (4000 rpm for 10 minutes) was performed at room temperature (approximately 22°C).

Total RNA was extracted from plasma samples (200 µl) using the miRNeasy Serum/Plasma Kit (Qiagen) according to manufacturer’s protocol. The samples were processed in random order with the researcher being blinded for the patient characteristics. The synthetic *Caenorhabditis elegans* miR-39-3p (cel-miR-39-3p) was added during the extraction procedure as an external reference RNA (1.6 × 10^8^ copies/μL). Since circulating lncRNA levels are generally low in plasma, a pre-amplification step was included using the iScript Explore One-Step RT and PreAmp kit (Biorad) according to manufacturer’s instruction. Briefly, 11.5 µL of the extracted RNA of each individual patient sample were used for the gDNA clearance reaction by mixing with 1.5 µL DNase reaction mix containing Dnase I and the buffer (included in the PreAmp kit). For preamplification and reverse transcription, 25 µL of PreAmp Supermix, 5 µL of assay primer pool, 5 µL of nuclease-free water, 1 µL of advanced reverse transcriptase and 1 µL of reaction booster were added to the DNase-treated RNA. Preamplification was done for 14 cycles in a thermocycler and the reactions diluted with 450 µL nuclease-free water and stored at −80°C.

Specific primers were generated following an intron-spanning design (min. size of the intron >700bp) to avoid false-amplification from any genomic DNA that might be present in the plasma samples. The generated primers were pre-tested using reverse transcribed RNA isolated from human embryonic kidney (HEK) cells. A single amplicon of the expected size was obtained as determined by semi-quantitative PCR and agarose gel electrophoresis. The amplicon was isolated from the gel and the sequence identity further confirmed by Sanger sequencing **(Supplemental Figure S2)**.

qPCR was performed, using the iQ SYBR Green Supermix kit by BioRad on a Viia7 Real-Time PCR system (Thermo Fisher Scientific) with the following cycling conditions: 15 min at 95°C, 40 cycles of 10 s at 95°C and 1 min at 60°C. For the determination of the external reference, the cel-miR-39-3p specific TaqMan miRNA assay (Applied Biosystems) was performed as previously described.^21^ The qPCR raw data was exported from the QuantStudioTM Real-Time PCR software and was analyzed using the LinRegPCR software.^22^ Cqs above 35 cycles were considered undetectable and were censored at the minimum level observed for the lncRNA. To ensure the optimal quality of the data, we analyzed spike-in RNA template to monitor the uniformity of the lncRNA quantification process. The relative expression was calculated as relative quantity (RQ): RQ= N0[lncRNA]/N0[cel-miR-39-3p]. RQ levels were log-transformed for statistical analyses.

### Statistical analysis

Statistical analyses were conducted using R software version 4.1.3 (the R Foundation for Statistical Computing, Vienna, Austria). Descriptive statistics were employed to summarize the characteristics of the study population, with continuous variables presented as the median (Quartile 1-Quartile 3) and categorical variables as the frequency (percentage). The demographic, clinical and biochemical characteristics were compared across PDE4DIPP6 tertiles using the Kruskal-Wallis test for continuous variables and Chi-squared test for categorical variables. Fisher’s exact test was employed for categorical variables, while the Mann–Whitney test was utilized for continuous variables in ACS and NSTEMI comparisons. To be considered confounder, a variable had to be associated with both ACS and PDE4DIPP6 level at p-value < 0.05. The correlation between continuous variables was assessed using Spearman’s rho coefficient.

To construct multivariable models, logistic regression was employed, with adjustments made for heparin use, considering its potential impact on lncRNA quantification.^23^ Receiver operating characteristic (ROC) curves were generated to estimate the area under the curve (AUC). Discrimination improvement between models, including hs-cTnT alone and hs-cTnT + PDE4DIPP6, was compared using the DeLong test ^24^. Reclassification ^25^ provided by the lncRNA was evaluated through the Integrated Discrimination Improvement (IDI) and Net Reclassification Improvement (NRI) indexes. ^23^

A significance level of p < 0.05 (two-tailed) was set for all analyses.

## Results

### Plasma levels of lncRNA PDE4DIPP6 are elevated in patients with ACS

**Table 1** details demographic, clinical and biochemical data categorized by ACS presence. ACS was diagnosed in 50.8% of patients. As anticipated, ACS patients were typically older, more frequently male and exhibited a substantial cardiovascular risk factor burden.

**Table 1.**
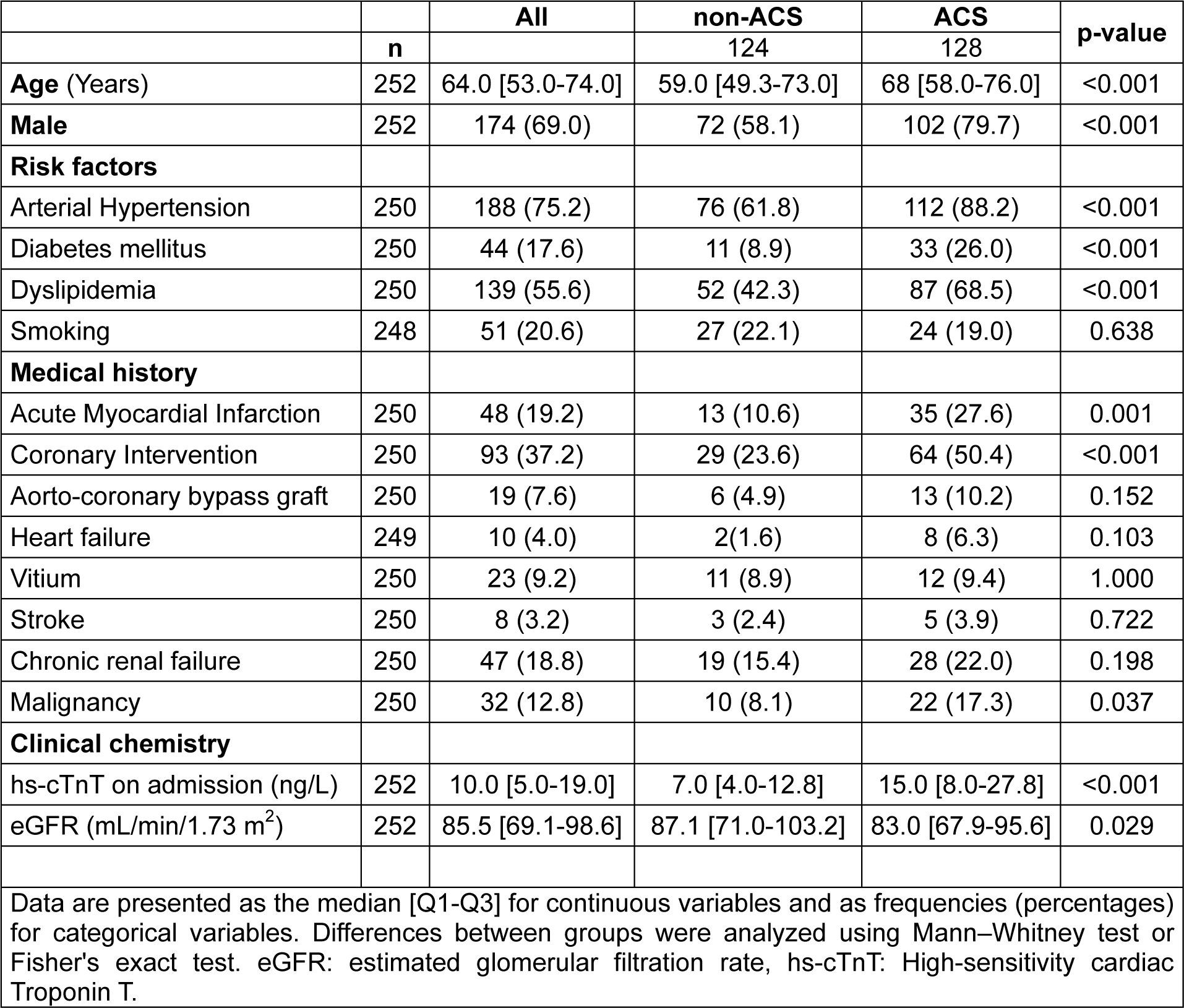
Characteristics of the patients stratified by the presence of acute coronary syndrome.

Elevated PDE4DIPP6 levels were observed in ACS patients compared to non-ACS individuals **(Figure 1)**. **Supplementary Table S1** provides comprehensive demographic, clinical, and biochemical data based on plasma PDE4DIPP6 levels. Tertiles of PDE4DIPP6 showed no differences in clinical characteristics or comorbidities, suggesting that the lncRNA levels were not associated with any confounder. Moreover, no significant correlation between circulating PDE4DIPP6 and hs-cTnT levels was found (rho=0.071) **(Table 2)**, implying a mechanism beyond myocardial necrosis and suggesting potential added diagnostic value to hs-cTnT. To test this hypothesis, we assessed the combined diagnostic potential of both hs-cTnT and PDE4DIPP6. As shown on **Table 3**, the addition of PDE4DIPP6 to a model containing hs-cTnT enhanced ACS discrimination (AUC hs-TnT + PDE4DIPP6 = 0.734; p-value = 0.003) and allowed significant patient reclassification with IDI = 0.066 (p-value < 0.001) and NRI = 0.471 (p-value < 0.001).

**Figure 1.**
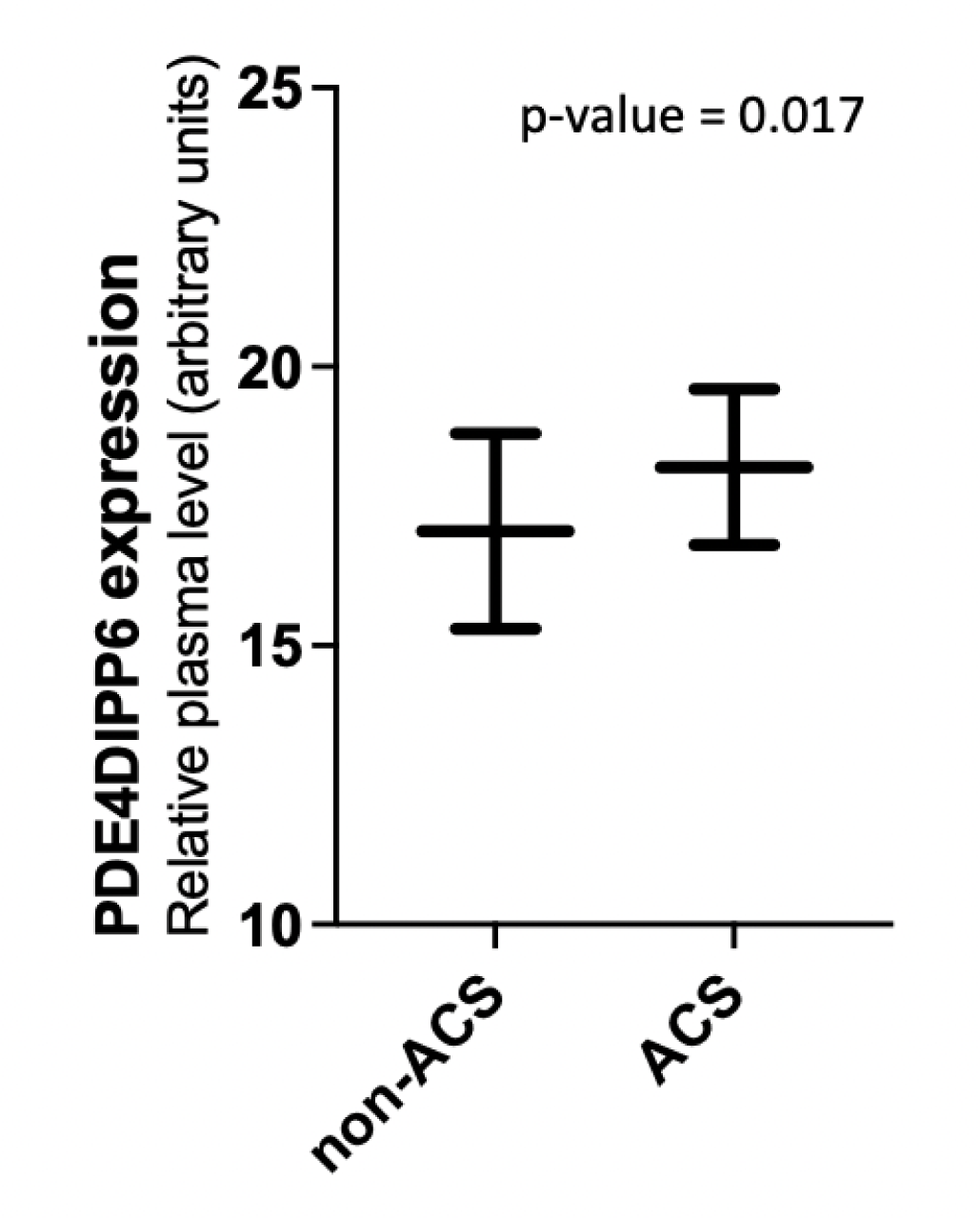
PDE4DIPP6 levels in patients with and without ACS. PDE4DIPP6 level was assessed in plasma using RT-qPCR. Relative quantification was performed using cel-miR-39-3p for normalization. Relative expression levels were log-transformed for statistical analyses. Differences between groups were analysed using Mann–Whitney test. The results are shown as the median with whiskers to minimum and maximum. P-value describes the significance level.

**Table 2.** Correlation between PDE4DIPP6 and hs-cTnT levels.

|  | **Spearman's rho** | **p-value** |
| --- | --- | --- |
| **hs-cTnT (ng/L)** | 0.071 | 0.324 |

Results are presented using the Spearman's rho. hs-cTnT: High-sensitivity Troponin T.

**Table 3.** Performance of circulating PDE4DIPP6 as a biomarker of acute coronary syndrome.

|  | Discrimination |  | Reclassification |  |  |  |
| --- | --- | --- | --- | --- | --- | --- |
|  | AUC | p-value (vs hs-cTnT) | IDI | p-value | NRI | p-value |
| **hs-cTnT (cutoff 5 ng/L)** | 0.650 | NA | Reference model | NA | Reference model | NA |
| **hs-cTnT (cutoff 5 ng/L) + PDE4DIPP6** | 0.734 | 0.003 | 0.066 | <0.001 | 0.471 | <0.001 |

AUC: Area under the curve; hs-cTnT: High-sensitivity Troponin T; IDI: Integrated discrimination improvement index; NRI: Continuous net reclassification improvement index. NA: Not applicable.

### Performance of circulating PDE4DIPP6 as a biomarker for NSTEMI

The discovery that PDE4DIPP6 enhances the differentiation between patients with and without ACS is highly promising. However, given that ACS is an umbrella term encompassing various clinical presentations requiring distinct treatments, the clinical utility of this analysis is constrained. We therefore further explored the potential of circulating PDE4DIPP6 to discriminate between NSTEMI-diagnosed patients and those without NSTEMI. To do so, the 252 patient samples were stratified into those without NSTEMI and those with NSTEMI diagnosis (13.9%). **Table 4** summarizes baseline characteristics. As expected, NSTEMI patients were more likely male and had a higher cardiovascular risk factor burden.

**Table 4.**
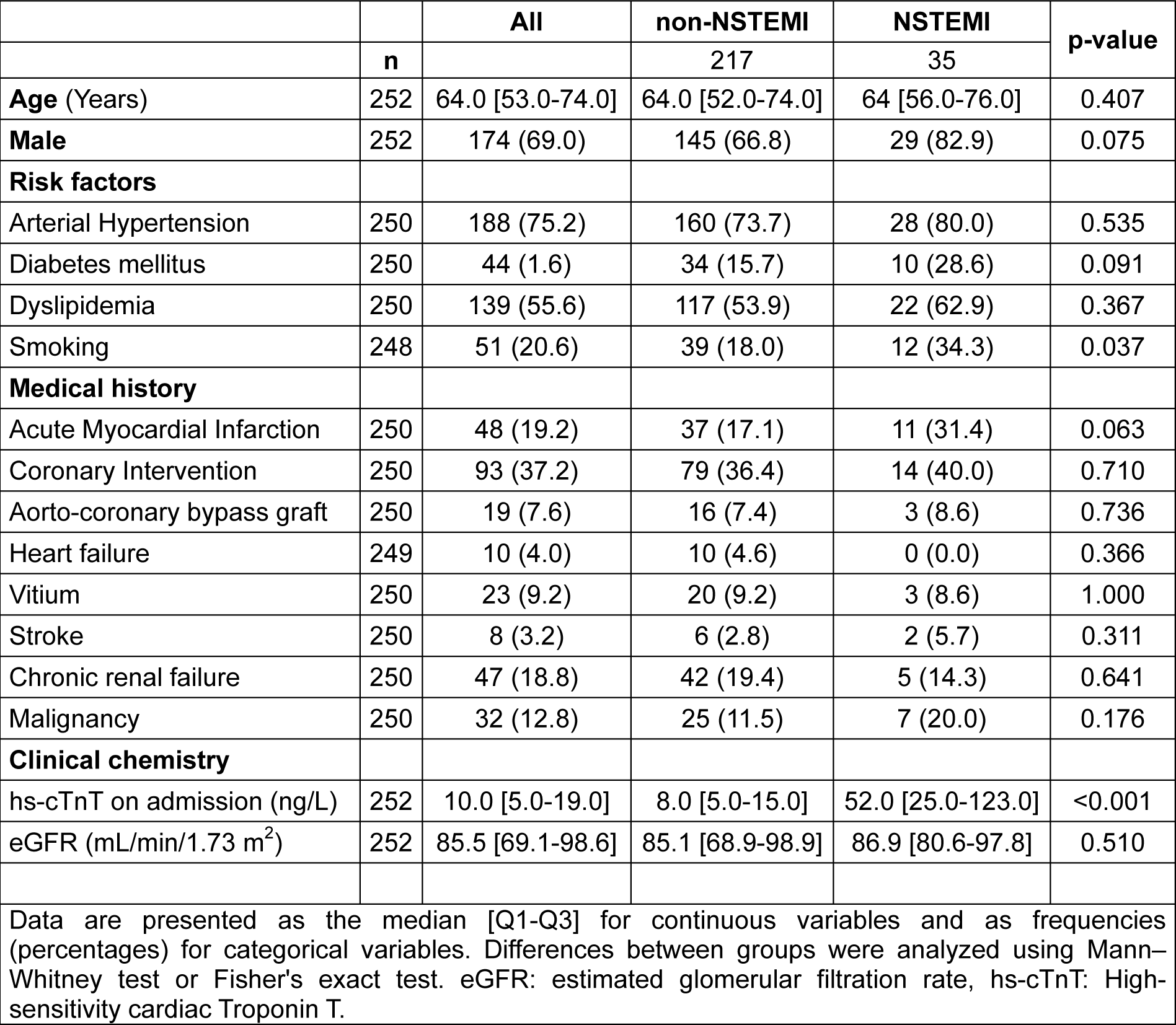
Characteristics of the patients stratified by the presence of NSTEMI.

|  |  | All | non-NSTEMI | NSTEMI | p-value |
| --- | --- | --- | --- | --- | --- |
|  | n |  | 217 | 35 |  |
| **Age** (Years) | 252 | 64.0 [53.0-74.0] | 64.0 [52.0-74.0] | 64 [56.0-76.0] | 0.407 |
| **Male** | 252 | 174 (69.0) | 145 (66.8) | 29 (82.9) | 0.075 |
| **Risk factors** |  |  |  |  |  |
| Arterial Hypertension | 250 | 188 (75.2) | 160 (73.7) | 28 (80.0) | 0.535 |
| Diabetes mellitus | 250 | 44 (1.6) | 34 (15.7) | 10 (28.6) | 0.091 |
| Dyslipidemia | 250 | 139 (55.6) | 117 (53.9) | 22 (62.9) | 0.367 |
| Smoking | 248 | 51 (20.6) | 39 (18.0) | 12 (34.3) | 0.037 |
| **Medical history** |  |  |  |  |  |
| Acute Myocardial Infarction | 250 | 48 (19.2) | 37 (17.1) | 11 (31.4) | 0.063 |
| Coronary Intervention | 250 | 93 (37.2) | 79 (36.4) | 14 (40.0) | 0.710 |
| Aorto-coronary bypass graft | 250 | 19 (7.6) | 16 (7.4) | 3 (8.6) | 0.736 |
| Heart failure | 249 | 10 (4.0) | 10 (4.6) | 0 (0.0) | 0.366 |
| Vitium | 250 | 23 (9.2) | 20 (9.2) | 3 (8.6) | 1.000 |
| Stroke | 250 | 8 (3.2) | 6 (2.8) | 2 (5.7) | 0.311 |
| Chronic renal failure | 250 | 47 (18.8) | 42 (19.4) | 5 (14.3) | 0.641 |
| Malignancy | 250 | 32 (12.8) | 25 (11.5) | 7 (20.0) | 0.176 |
| **Clinical chemistry** |  |  |  |  |  |
| hs-cTnT on admission (ng/L) | 252 | 10.0 [5.0-19.0] | 8.0 [5.0-15.0] | 52.0 [25.0-123.0] | <0.001 |
| eGFR (mL/min/1.73 m<sup>2</sup>) | 252 | 85.5 [69.1-98.6] | 85.1 [68.9-98.9] | 86.9 [80.6-97.8] | 0.510 |

Data are presented as the median [Q1-Q3] for continuous variables and as frequencies (percentages) for categorical variables. Differences between groups were analyzed using Mann–Whitney test or Fisher's exact test. eGFR: estimated glomerular filtration rate, hs-cTnT: High-sensitivity cardiac Troponin T.

Figure 2 illustrates the higher circulating PDE4DIPP6 levels in NSTEMI patients compared to those without NSTEMI. The addition of PDE4DIPP6 significantly increased the discriminative power of hs-cTnT (AUC = 0.944, p=0.003) **(Table 5)**. Additionally, PDE4DIPP6 significantly facilitated patient reclassification when added to hs-cTnT, with IDI = 0.237 (p-value = 0.006) and NRI = 0.658 (p-value = 0.003) **(Table 5)**.

**Figure 2.**
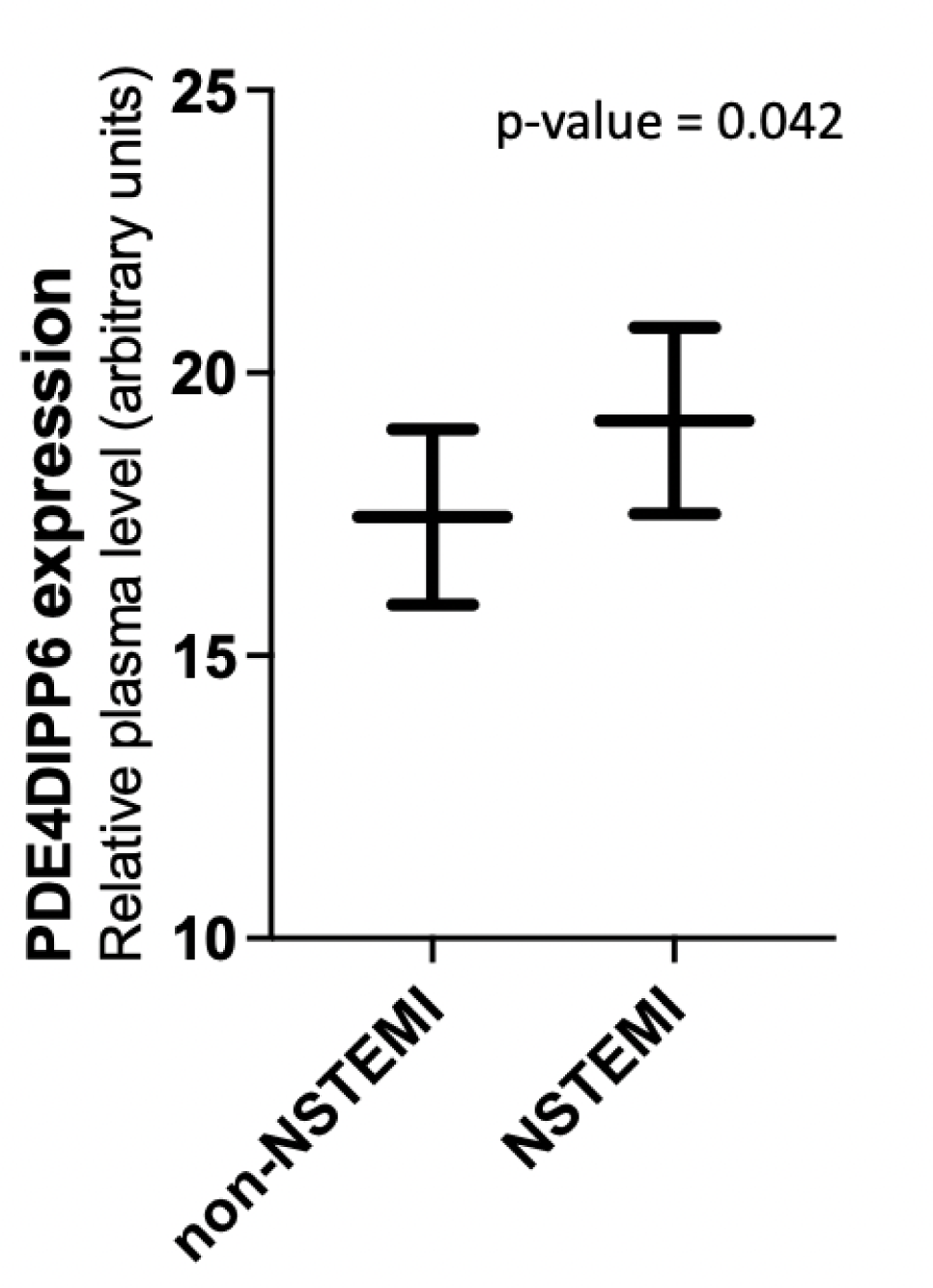
PDE4DIPP6 levels in patients with and without NSTEMI. PDE4DIPP6 level was assessed in plasma using RT-qPCR. Relative quantification was performed using cel-miR-39-3p for normalization. Relative expression levels were log-transformed for statistical analyses. Differences between groups were analysed using Mann–Whitney test. The results are shown as the median with whiskers to minimum and maximum. P-value describes the significance level.

**Table 5.** Performance of circulating PDE4DIPP6 as a biomarker of NSTEMI.

|  | Discrimination |  | Reclassification |  |  |  |
| --- | --- | --- | --- | --- | --- | --- |
|  | AUC | p-value (vs hs-cTnT) | IDI | p-value | NRI | p-value |
| hs-cTnT (cutoff 14 ng/L + change) | 0.859 | NA | Reference model | NA | Reference model | NA |
| hs-cTnT (cutoff 14 ng/L + change) + PDE4DIPP6 | 0.944 | 0.003 | 0.237 | 0.006 | 0.658 | 0.003 |

AUC: Area under the curve; hs-cTnT: High-sensitivity Troponin T; IDI: Integrated discrimination improvement index; NRI: Continuous net reclassification improvement index. NA: Not applicable.

## Discussion

To date, the cardiac enzymes Troponin I and T are acknowledged as the gold standard biomarkers for managing AMI.^26^ In clinical practice, there is no imperative need for a new biomarker to entirely supplant cardiac Troponin T, given its already established reliability in AMI. However, implementation of hs-cTnT has impacted the specificity of diagnostic power, leading to occasional false diagnoses.^6^ The primary objective of this study was to identify a novel circulating lncRNAs that could enhance medical decision-making within the context of ACS. More specifically, the aim was to define a lncRNA capable of augmenting the information provided by hs-cTnT in the clinical management of NSTEMI.

Initially, we reported elevated levels of PDE4DIPP6 in the plasma of ACS patients compared to those without ACS. Moreover, no correlation was observed between circulating PDE4DIPP6 and hs-cTnT levels, despite both being upregulated in ACS and NSTEMI. No other associations were noted between lncRNA levels and potential confounding factors, such as patient characteristics and medical history. Based on these data, we evaluated the potential of PDE4DIPP6 to complement the diagnostic value of hs-cTnT. Our findings indicated that the lncRNA increased the discrimination value of hs-cTnT and correctly reclassified misclassified patients not only in ACS but also in NSTEMI. Therefore, current results indicate that PDE4DIPP6 provides additional diagnostic insights beyond hs-cTnT, suggesting its potential to improve the clinical management of ACS. This holds true even within a patient population characterized by a diverse range of symptoms and a substantial prevalence of comorbidities.

While comparisons with previous studies on lncRNAs and ACS are limited by differences in experimental designs, study populations, sample matrix and candidates evaluated, these findings support the notion that lncRNAs could be valuable in the management of AMI. Indeed, Vasourt et al.^27^ demonstrated that patients with STEMI had lower levels of the lncRNAs ANRIL, KCNQ1OT1, MIAT and MALAT1 when compared with patients with NSTEMI. In this line, our group have previously reported that the levels of circulating LIPCAR are altered during the early stage of post-AMI left ventricle remodeling and constitute a prognostic indicator of chronic heart failure.^28^ More recently, other authors have proposed that diverse lncRNAs; ENST00000444488.1,^29^ HIF1A-AS2,^30^ TTTY15 and HULC,^31^ could also contribute to the diagnosis of AMI. Overall, and similar to observations with other noncoding RNAs,^32,33^ our results support a significant opportunity to enhance clinical assessment and guide decision-making using a model based on readily available clinical data and noncoding transcripts. Future studies using global lncRNA profiling are fundamental to identify novel candidates with potential as biomarkers in ACS.

Previous evidence suggests that lncRNAs may function as mediators in cell-to-cell communication.^10^ Hence, a pivotal question arises concerning the biological relevance of the observed alteration in circulating PDE4DIPP6 levels and, consequently, its potential causal involvement in ACS. The identified association between ACS and circulating levels of PDE4DIPP6 implies a conceivable link between this lncRNA and molecular mechanisms implicated in the pathology of ACS. The information provided by PDE4DIPP6 may offer complementary insights to available health records, potentially reflecting disease-associated mechanisms not captured by conventional markers such as hs-cTnT levels or cardiovascular risk factors. Additionally, our data provide insights into a potential cellular origin of circulating PDE4DIPP6. It is well known that PDE4 contributes to the regulation of the second messenger cAMP in cardiomyocytes. In turn, cAMP activates the cAMP-dependent protein kinase (PKA), which phosphorylates specific protein, resulting in the initiation of a variety of signals.^34,35^ This includes the activation of the sympathetic nervous system and therefore the myocardial constriction.^36^ For example, the phosphodiesterase 4D3 (PDE4D3) was found in the cardiac ryanodine receptor (RyR2)/calcium-release-channel complex, which is the major Ca2+-release channel required for excitation-contraction coupling in the heart muscle.^34^ Also, certain PDE isoforms co-localize with PKA as part of negative feedback mechanisms which may protect from excess beta-adreno-receptor stimulation of calcium transporters, during cardiac excitation-contraction coupling.^37^ Various studies suggest that a reduction in the PDE4D activity contributes to heart failure, cardiomyopathy and other heart conditions.^33,35,36^ It is essential to mention that our study design does not permit the determination of causal relationships. While the observed associations provide valuable insights into potential connections, further research is warranted to elucidate the intricate molecular dynamics within the context of ACS. Specifically, additional *in vitro* and *in vivo* studies, with a special focus on both hiPSC-CM and HCF, are required for a more comprehensive understanding of the underlying mechanisms.

The strengths of this study lie in its real-world setting, including the assessment of PDE4DIPP6 alongside electronic health records and concurrent tests as well as the inclusion of patients with a broad spectrum of symptoms [with the median age of 64 years and 69 % male sex, similar to and therefore representative of the entire original cohort (age median 65 years and 66% male^38^)]. The accessibility of samples for PDE4DIPP6 analysis could facilitate its implementation in routine care. However, although the results are promising, certain limitations that warrant further investigation should be noted. First, the need for validation in larger, independent populations is imperative to affirm the robustness and generalizability of our findings. Second, we cannot exclude the effect of diseases or conditions that were not recorded in this study and that may influence plasma PDE4DIPP6 levels. Third, the measurement of PDE4DIPP6 itself poses challenges, necessitating careful consideration of both pre-analytical and analytical factors. Specifically, the preamplification process presents a hurdle.^39^ It is crucial to note that the normalization process utilizing a transcript, such as cel-miR-39-3p, which possesses distinct biochemical properties, may introduce potential impacts on data analysis. However, to date, there is a notable absence of a normalization strategy universally acknowledged as the gold standard. The choice of normalization method remains a subject of ongoing exploration and discussion within the scientific community.^39^ It is noteworthy that heparin administration is prevalent in ACS patients. Consequently, we incorporated the administration of heparin or fondaparinux into the multivariate model to account for this potential source of bias in lncRNA quantification.^23^ Forth, our study focused on assessing the biomarker potential of a specific lncRNA candidate. The selection of PDE4DIPP6 was informed by prior results and the collective expertise of our research group. In conclusion, our results suggest that PDE4DIPP6 could serve as a biomarker to improve medical decision making in ACS, particularly in NSTEMI. Further investigations are warranted to explore specific pathways related to PDE4DIPP6, connecting its dynamic regulation to ACS and, especially, NSTEMI.

## Abbreviations

**RT**: Reverse transcription reaction with Reverse transcriptase; **95% CI**: 95% confidence interval; **ACS**: Acute coronary syndrome; **AMI**: Acute myocardial infarction; **AUC**: Area under the curve; **C. elegans**: Caenorhabditis elegans; **HCF**: human cardiac fibroblasts; **HEK**: Human embryonic kidney; **hiPSC-CM**: Human induced pluripotent stem cell-derived cardiomyocytes; **ICD:** Implantable Cardioverter-Defibrillator; **IDI:** Integrated discrimination improvement index; **lncRNA**: Long noncoding ribonucleic acid; **NRI**: Continuous net reclassification improvement index; **NSTEMI**: Non-ST-elevation myocardial infarction; **nt**: Nucleotides; **ROC**: Receiver operating characteristic; **RQ**: Relative quantity; **STEMI**: ST-elevation myocardial infarction; **uAP**: Unstable angina pectoris

## Data Availability

All data is included in the manuscript

